# Radiographic Findings and other Predictors in Adults with Covid-19

**DOI:** 10.1101/2020.03.23.20041673

**Authors:** Kaiyan Li, Dian Chen, Shenchong Chen, Yuchen Feng, Chenli Chang, Zi Wang, Nan Wang, Guohua Zhen

## Abstract

As of March 20, 2020, there were 234,073 confirmed cases of coronavirus disease 2019 (Covid-19) and 9,840 deaths worldwide ^1^. Older age and elevated d-dimer are reported risk factors for Covid-19 ^2,3^. However, whether early radiographic change is a predictor of fatality remains unknown. We retrospectively reviewed records of all laboratory-confirmed patients admitted to a quarantine unit at Tongji Hospital, a large regional hospital in Wuhan, China, between January 31 and March 5, 2020. The Tongji Hospital ethics committee approved this study.

---

A total of 128 patients were admitted. 102 patients were confirmed to have severe acute respiratory syndrome coronavirus 2 (SARS-CoV-2) infection using RNA detection. As of March 20, 82 confirmed patients were discharged, 15 died, and 5 remained hospitalized. The median age was 57 years (range, 27 - 85), 59 (58%) were male, and 44 (43%) patients had a comorbidity. The most common symptoms were fever, cough, and dyspnea (Table S1). When compared with survivors, non-survivors were older and more likely to have lymphopenia, elevated lactate dehydrogenase (LDH), elevated d-dimer, and increased hypersensitive troponin I (Table S1, S2). In a multivariate regression model that included these predictors, older age and elevated LDH were independent risk factors for fatality (Table S3).

Twenty-one survivors and 11 non-survivors had CT scans within the first week. We used severity score to quantify the extent of lung opacification as described in the Supplementary Appendix. The total severity score and number of involved lung lobes within the first week were significantly greater in non-survivors compared to survivors (Table S4). Using univariate logistic regression analysis, higher total severity score (≥15) (odds ratio 53, 95% CI 3-369; p = 0.003), and more involved lung lobes (5 involved lobes) (9, 2-53; p = 0.016) in CT images within the first week were significantly associated with fatality (Figure 1A B, Table S5). Moreover, in this subset of patients with CT data within the first week, higher total severity score was the only independent risk factor in a multivariate analysis incorporated the predictors discussed above (older age, lymphocytopenia, elevated LDH, elevated d-dimer, and increased troponin I) (Table S5). For survivors with serial CT scans performed over four weeks, total severity score peaked in the second week (Figure 1C, Table S4).

**Figure 1.**
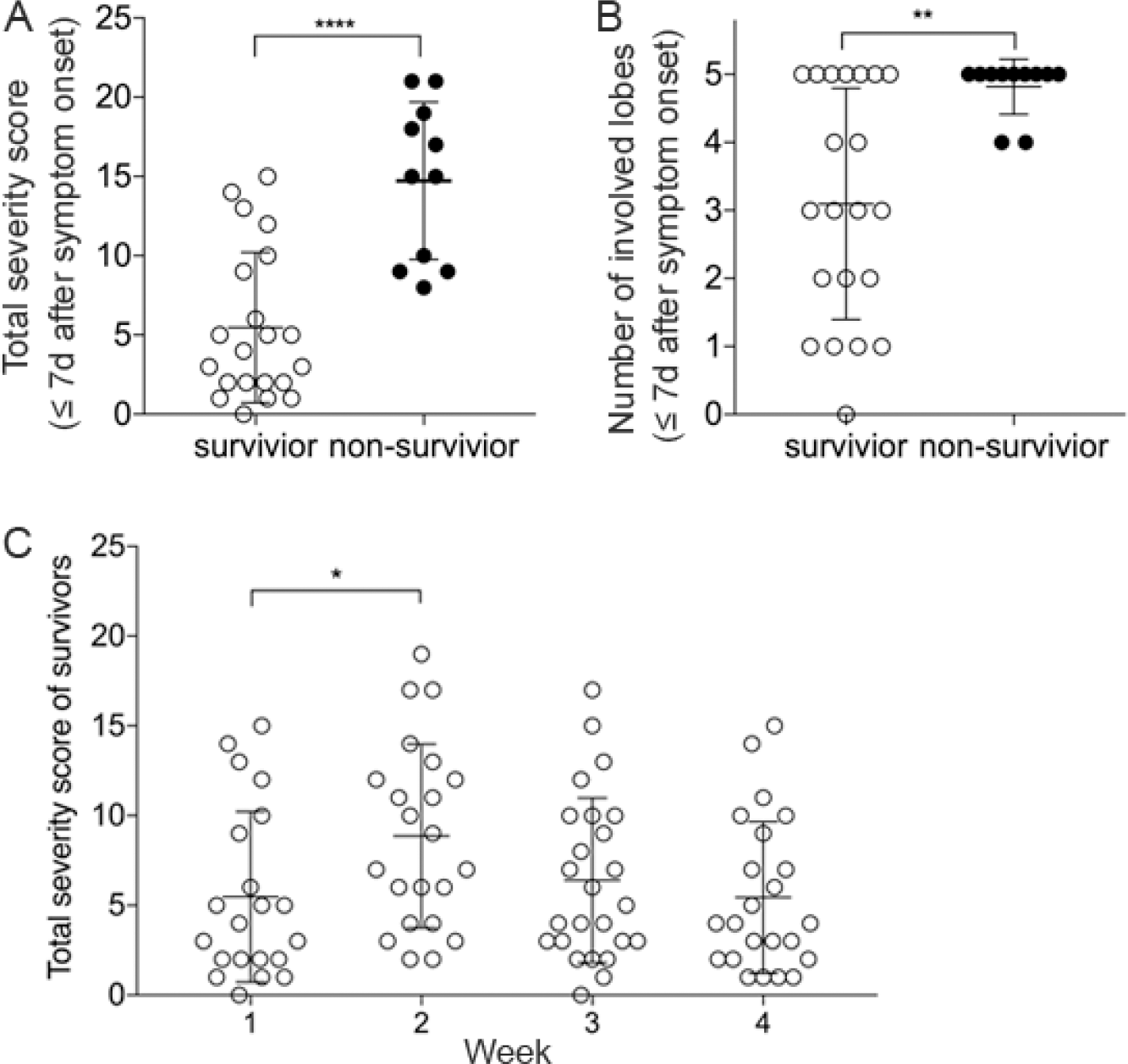
Total severity score and the number of involved lung lobes in CT images. **A-B**, total severity score (A) and the number of involved lung lobes (B) for CT images of survivors and non-survivors within the first week (≤7d) after symptom onset. **C**, total severity score for CT images of survivors over the 4 weeks after symptom onset. Values of survivors and non-survivors are presented with open and closed circles, respectively. Mann-Whitney U test was used to compare the values of survivors in Week 1 with those of non-survivors, and with those of survivors in Week 2, 3, 4, respectively. ****, *p* < 0.0001; **, *p* < 0.01; *, *p* < 0.05.

This report suggests that the extent of lung lesions in early CT images is a potential predictor of poor outcome of Covid-19. This will help clinicians to identify the patients with poor prognosis at early stage.

## Data Availability

NA

## Supplementary Appendix

### Methods

#### Study Population and Data Collection

This retrospective study was approved by the institutional ethics board of Tongji Hospital of Huazhong University of Science and Technology. Written informed consent was waived. The study included all patients with laboratory-confirmed COVID-19 admitted to a quarantine unit of Tongji Hospital, a large regional hospital in Wuhan, China, between January 31 and March 5, 2020. COVID-19 patients were diagnosed according to World Health Organization (WHO) interim guideline ^1^. Confirmed cases were defined by the positive findings in reverse-transcriptase–polymerase-chain-reaction (RT-PCR) assay of throat swab specimens. Clinical characteristics, laboratory test results, and treatment information were extracted from electronic medical records. All laboratory testing and radiological examination were performed according to the clinical care needs of the patient.

#### RT-PCR for SARS-CoV-2

Throat swab specimens were tested for SARS-CoV-2 using real-time RT-PCR according to the WHO protocol ^3^. The following primers and probes were used for real-time RT-PCR detection of N gene of SARS-CoV-2: N forward primer 5’-GAGCCTTGAATACACCAAAAG-3’, N reverse primer 5’-GCACGATTGCAGCATTGTTAGCAGGATT-3’, N probe 5’-FAMCACATTGGCACCCGCAATCC-MGB-3’. Positive results were confirmed in two independent real-time RT-PCR assays.

#### Chest CT Protocols and Evaluation

High-resolution transverse CT images were obtained using Optima 660 (GE Medical System, Milwaukee, USA) or Somatom Definition AS+ (Siemens Healthineers, Forchheim, Germany). Tube voltage was 100 or 120 kV, and automatic tube current modulation was 100 - 400 mA. All images were reconstructed with a slice thickness of 1.0mm or 1.25mm. The CT images were reviewed by two radiologists (ZW and NW) who were blinded to the final outcome of the patients. Images were reviewed independently. Any disagreements were resolved by discussion and consensus.

A scoring system was used to estimate the extent of lung opacification based on the area involved ^2^. Each of the five lung lobes was visually scored from 0 to 5 as: 0, no involvement; 1, < 5% involvement; 2, 5% - 25% involvement; 3, 26% - 49% involvement; 4, 50% - 75% involvement; 5, > 75% involvement. The total severity score was the sum of scores of each lobe, ranging from 0 (no involvement) to 25 (maximum involvement).

#### Statistical Analysis

Statistical analysis was done with SPSS Statistics Software (version 26; IBM, New York, USA). Continuous variables were presented as median (IQR) and analyzed using Mann-Whitney U test; categorical variables were presented as number (%) and analyzed by χ^2^ test or Fisher’s exact test between survivors and non-survivors where appropriate. Univariable and multivariable logistic regression models were used to estimate odds ratios and the 95% confidence intervals of the risk factors associated with fatal outcome. A two-sided α of less than 0.05 was considered statistically significant.

**Table S1.**
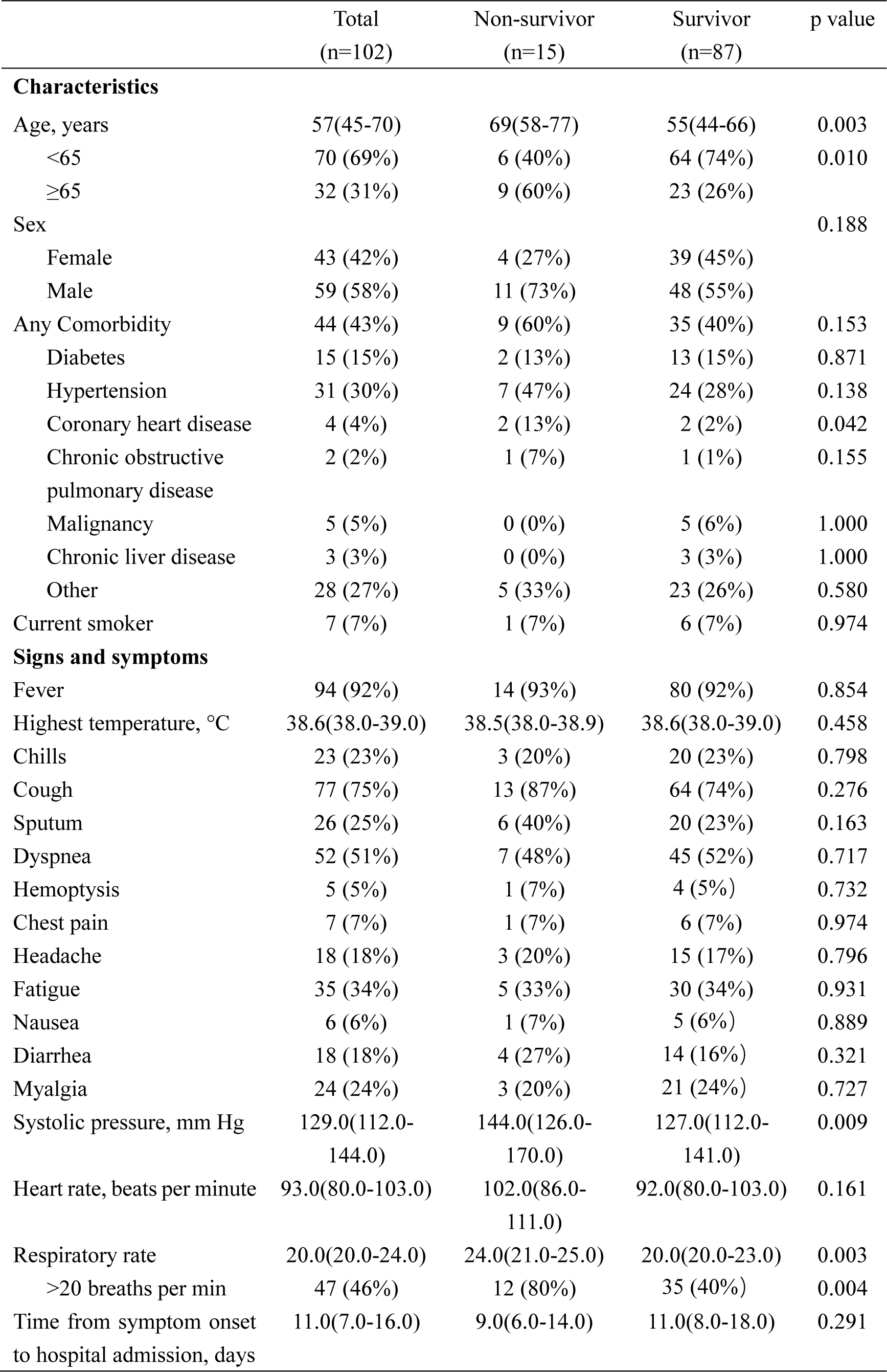
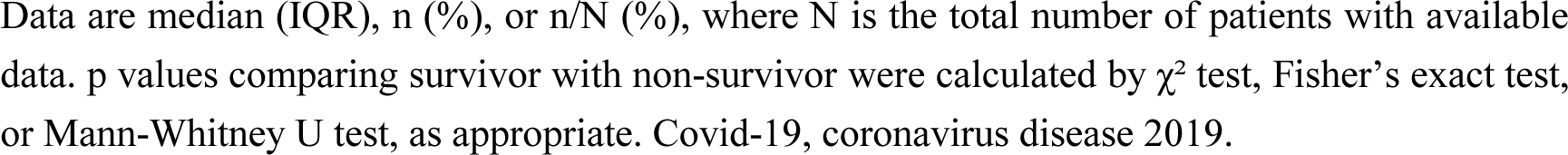
Demographics and baseline characteristics of patients with Covid-19.

|  | Total<br>(n=102) | Non-survivor<br>(n=15) | Survivor<br>(n=87) | p value |
| --- | --- | --- | --- | --- |
| **Characteristics** |  |  |  |  |
| Age, years | 57(45-70) | 69(58-77) | 55(44-66) | 0.003 |
| <65 | 70 (69%) | 6 (40%) | 64 (74%) | 0.010 |
| ≥65 | 32 (31%) | 9 (60%) | 23 (26%) |  |
| Sex |  |  |  | 0.188 |
| Female | 43 (42%) | 4 (27%) | 39 (45%) |  |
| Male | 59 (58%) | 11 (73%) | 48 (55%) |  |
| Any Comorbidity | 44 (43%) | 9 (60%) | 35 (40%) | 0.153 |
| Diabetes | 15 (15%) | 2 (13%) | 13 (15%) | 0.871 |
| Hypertension | 31 (30%) | 7 (47%) | 24 (28%) | 0.138 |
| Coronary heart disease | 4 (4%) | 2 (13%) | 2 (2%) | 0.042 |
| Chronic obstructive<br>pulmonary disease | 2 (2%) | 1 (7%) | 1 (1%) | 0.155 |
| Malignancy | 5 (5%) | 0 (0%) | 5 (6%) | 1.000 |
| Chronic liver disease | 3 (3%) | 0 (0%) | 3 (3%) | 1.000 |
| Other | 28 (27%) | 5 (33%) | 23 (26%) | 0.580 |
| Current smoker | 7 (7%) | 1 (7%) | 6 (7%) | 0.974 |
| **Signs and symptoms** |  |  |  |  |
| Fever | 94 (92%) | 14 (93%) | 80 (92%) | 0.854 |
| Highest temperature, °C | 38.6(38.0-39.0) | 38.5(38.0-38.9) | 38.6(38.0-39.0) | 0.458 |
| Chills | 23 (23%) | 3 (20%) | 20 (23%) | 0.798 |
| Cough | 77 (75%) | 13 (87%) | 64 (74%) | 0.276 |
| Sputum | 26 (25%) | 6 (40%) | 20 (23%) | 0.163 |
| Dyspnea | 52 (51%) | 7 (48%) | 45 (52%) | 0.717 |
| Hemoptysis | 5 (5%) | 1 (7%) | 4 (5%) | 0.732 |
| Chest pain | 7 (7%) | 1 (7%) | 6 (7%) | 0.974 |
| Headache | 18 (18%) | 3 (20%) | 15 (17%) | 0.796 |
| Fatigue | 35 (34%) | 5 (33%) | 30 (34%) | 0.931 |
| Nausea | 6 (6%) | 1 (7%) | 5 (6%) | 0.889 |
| Diarrhea | 18 (18%) | 4 (27%) | 14 (16%) | 0.321 |
| Myalgia | 24 (24%) | 3 (20%) | 21 (24%) | 0.727 |
| Systolic pressure, mm Hg | 129.0(112.0-144.0) | 144.0(126.0-170.0) | 127.0(112.0-141.0) | 0.009 |
| Heart rate, beats per minute | 93.0(80.0-103.0) | 102.0(86.0-111.0) | 92.0(80.0-103.0) | 0.161 |
| Respiratory rate | 20.0(20.0-24.0) | 24.0(21.0-25.0) | 20.0(20.0-23.0) | 0.003 |
| >20 breaths per min | 47 (46%) | 12 (80%) | 35 (40%) | 0.004 |
| Time from symptom onset<br>to hospital admission, days | 11.0(7.0-16.0) | 9.0(6.0-14.0) | 11.0(8.0-18.0) | 0.291 |

Data are median (IQR), n (%), or n/N (%), where N is the total number of patients with available data. p values comparing survivor with non-survivor were calculated by $\chi^2$ test, Fisher's exact test, or Mann-Whitney U test, as appropriate. Covid-19, coronavirus disease 2019.

**Table S2.**
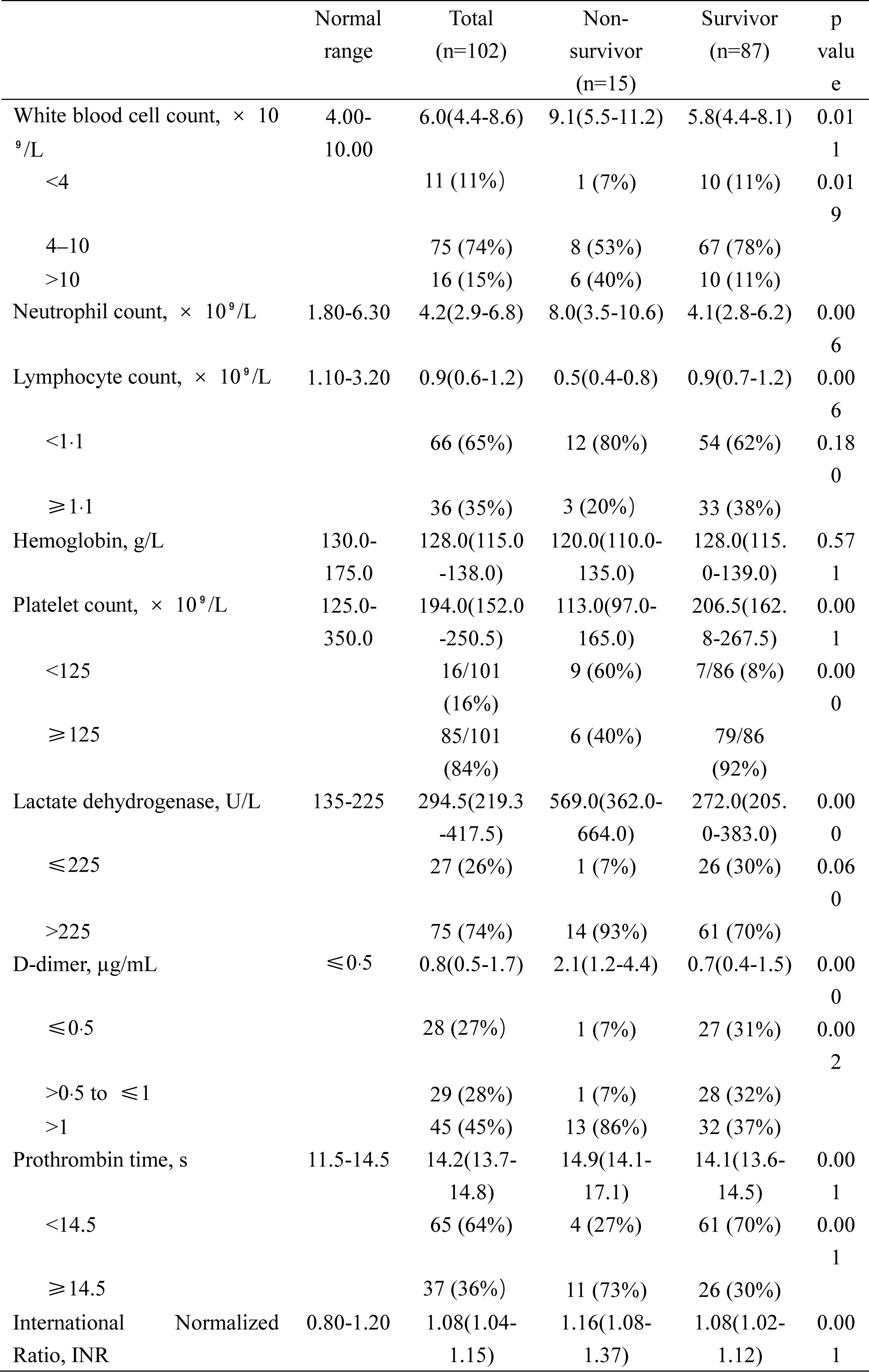
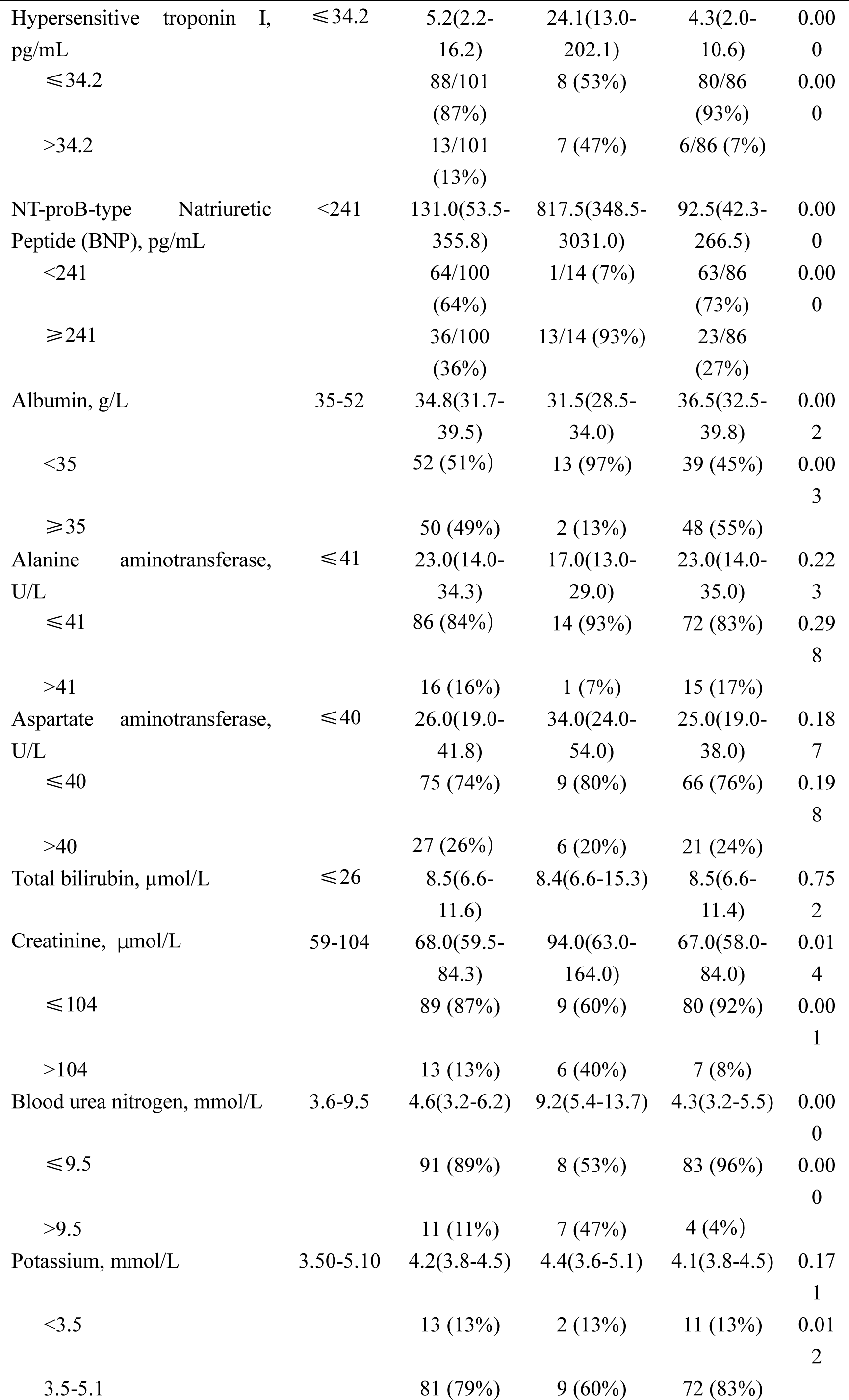
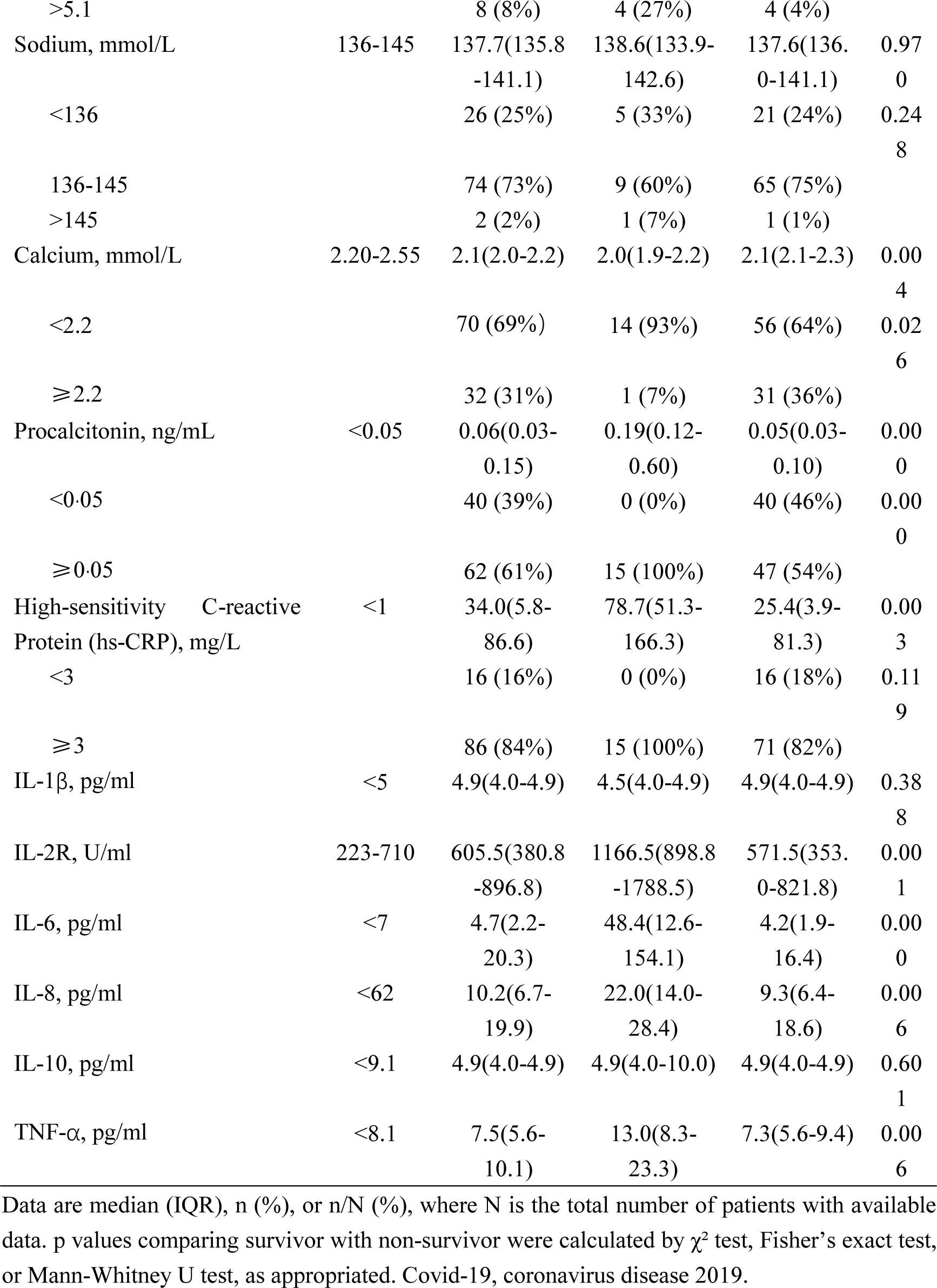
Laboratory findings of patients with Covid-19 on admission.

**Table S3.** Risk factors associated with fatality.

|  | Univariable<br>OR (95% CI) | p value | Multivariable<br>OR (95% CI) | p value |
| --- | --- | --- | --- | --- |
| **Demographics and clinical characteristics** |  |  |  |  |
| Age, years* | 1.07(1.02-1.11) | 0.006 | 1.063(1.006-1.124) | 0.031 |
| <65 | 1 (ref) | .. | .. | .. |
| ≥65 | 4.17(1.34-13.02) | 0.014 | .. | .. |
| Female sex (vs male) | 0.45(0.13-1.52) | 0.196 | .. | .. |
| Comorbidity present<br>(vs not present) |  |  |  |  |
| Diabetes | 0.88(0.18-4.34) | 0.871 | .. | .. |
| Hypertension | 2.30(0.75-7.03) | 0.120 | .. | .. |
| Coronary heart disease | 6.54(0.85-50.54) | 0.072 | .. | .. |
| Other | 1.39(0.43-4.50) | 0.582 | .. | .. |
| Current smoker | 0.96(0.11-8.63) | 0.974 |  |  |
| **Laboratory findings** |  |  |  |  |
| White blood cell count,<br>× 10<sup>9</sup>/L | 1.29(1.07-1.56) | 0.008 | .. | .. |
| <4 | 0.84(0.09-7.43) | 0.873 | .. | .. |
| 4-10 | 1 (ref) | .. | .. | .. |
| >10 | 5.03(1.44-17.54) | 0.011 | .. | .. |
| Neutrophil count, × 10<sup>9</sup>/L* | 1.33(1.11-1.59) | 0.002 | .. | .. |
| Lymphocyte count, × 10<sup>9</sup>/L* | 0.17(0.04-0.79) | 0.024 | .. | .. |
| Platelet count, × 10<sup>9</sup>/L | 0.98(0.97-0.99) | 0.001 | .. | .. |
| <125 | 16.93(4.66-61.51) | 0.000 | .. | .. |
| ≥125 | 1 (ref) | .. | .. | .. |
| Lactate dehydrogenase,<br>U/L* | 1.01(1.00-1.02) | 0.000 | 1.010(1.005-1.015) | 0.000 |
| ≤225 | 1 (ref) | .. | .. | .. |
| >225 | 5.97(0.75-47.77) | 0.092 | .. | .. |
| D-dimer, µg/mL | 1.14(1.04-1.25) | 0.007 | .. | .. |
| ≤0.5 | 1 (ref) | .. | .. | .. |
| >0.5 to ≤1 | 0.96(0.06-16.21) | 0.980 | .. | .. |
| >1 | 10.97(1.35-89.34) | 0.025 | .. | .. |
| Prothrombin time, s | 1.08(0.96-1.21) | 0.210 | .. | .. |
| <14.5 | 1 (ref) | .. | .. | .. |
| ≥14.5 | 6.45(1.88-22.14) | 0.003 | .. | .. |
| Hypersensitive troponin I,<br>pg/mL | 1.00(0.99-1.01) | 0.313 | .. | .. |
| ≤34.2 | 1 (ref) | .. | .. | .. |
| >34.2 | 11.67(3.15-43.26) | 0.000 | .. | .. |
| NT-proB-type natriuretic peptide (BNP), pg/mL | 1.00(1.00-1.00) | 0.706 | .. | .. |
| <241 | 1 (ref) | .. | .. | .. |
| ≥241 | 35.61(4.41-287.68) | 0.001 | .. | .. |
| Albumin, g/L | 0.82(0.71-0.93) | 0.003 | .. | .. |
| <35 | 8.00(1.70-37.60) | 0.008 | .. | .. |
| ≥35 | 1 (ref) | .. | .. | .. |
| Alanine aminotransferase, U/L | 0.97(0.93-1.02) | 0.204 | .. | .. |
| ≤41 | 1 (ref) | .. | .. | .. |
| >41 | 0.34(0.04-2.81) | 0.319 | .. | .. |
| Creatinine, μmol/L | 1.00(0.99-1.01) | 0.535 | .. | .. |
| ≤104 | 1 (ref) | .. | .. | .. |
| >104 | 7.62(2.10-27.68) | 0.002 | .. | .. |
| Blood urea nitrogen, mmol/L | 1.12(1.02-1.23) | 0.018 | .. | .. |
| ≤9.5 | 1 (ref) | .. | .. | .. |
| >9.5 | 18.16(4.36-75.62) | 0.000 | .. | .. |
| Procalcitonin, ng/mL* | 1.86(0.82-4.20) | 0.138 | .. | .. |
| High-sensitivity C-reactive Protein (hs-CRP), mg/L* | 1.01(1.00-1.02) | 0.005 | .. | .. |
| IL-2R, U/ml* | 1.003(1.001-1.004) | 0.001 | .. | .. |
| IL-6, pg/ml* | 1.02(1.01-1.04) | 0.002 | .. | .. |
| TNF-α, pg/ml* | 1.19(1.06-1.34) | 0.005 | .. | .. |

OR=odds ratio. \*Per 1 unit increase.

**Table S4.**
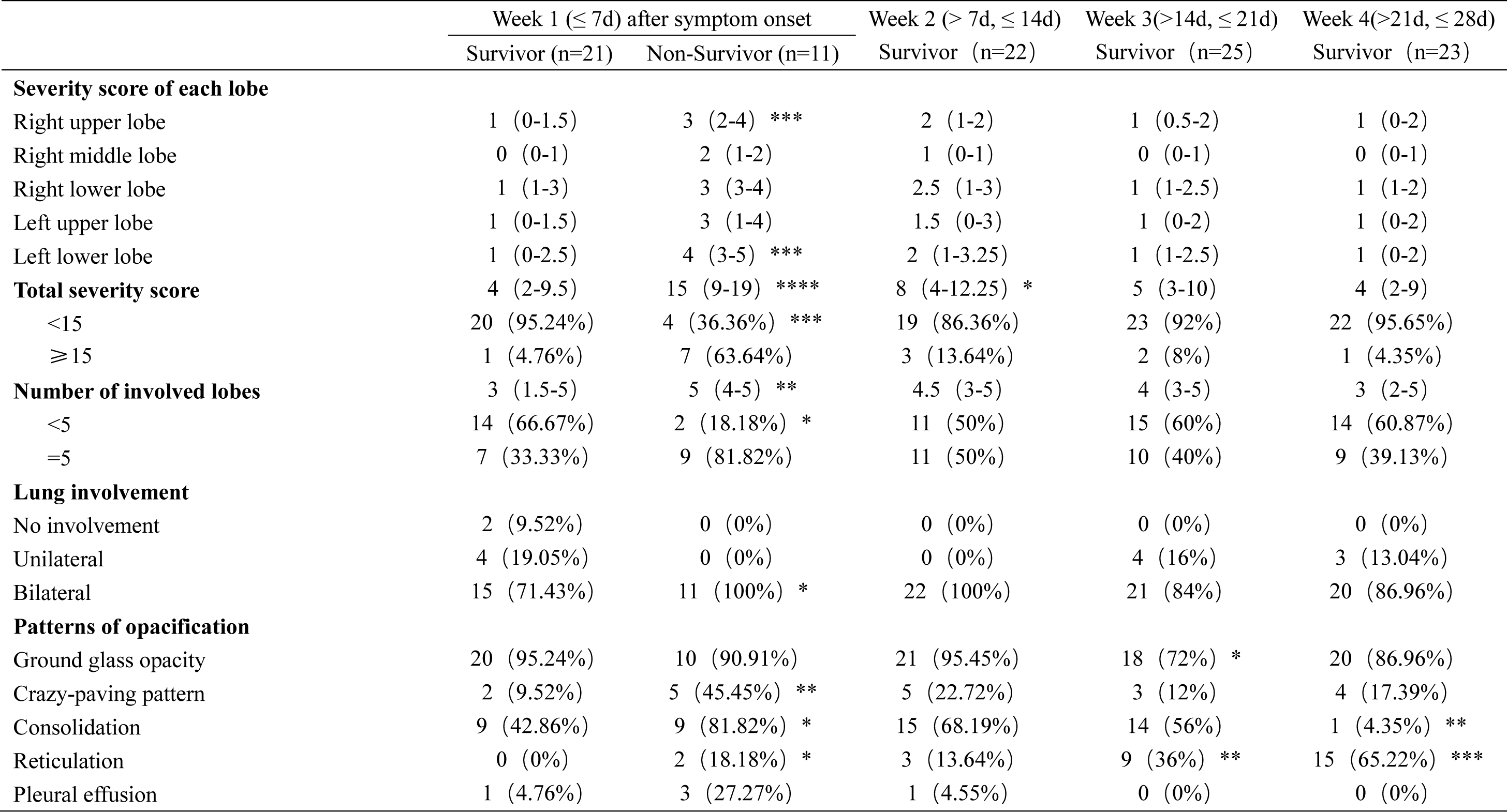
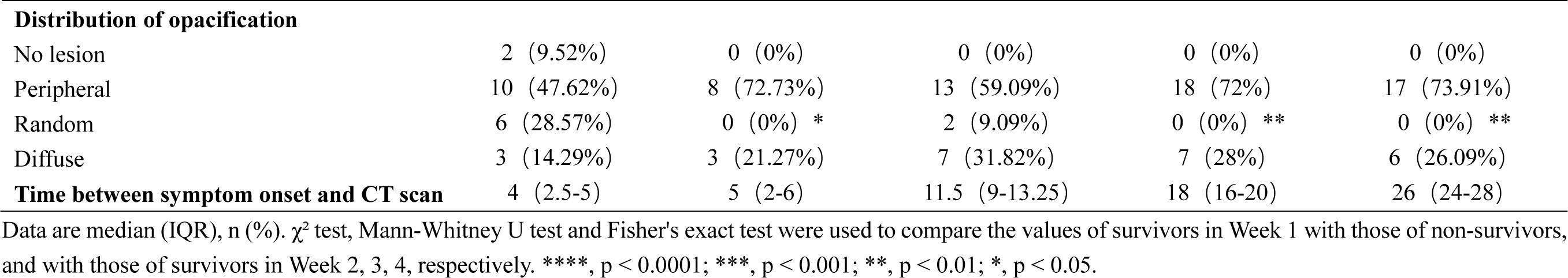
CT features of patients with Covid-19.

**Table S5.** Risk factors associated with fatality of the subset of patients with CT severity scores within the first week after symptom onset.

|  | Univariable<br>OR (95% CI) | p<br>value | Multivariable<br>OR (95% CI) | p<br>value |
| --- | --- | --- | --- | --- |
| **Demographics** |  |  |  |  |
| Age, years* | 1.06(1.01-1.13) | 0.027 | .. | .. |
| **Laboratory findings on admission** |  |  |  |  |
| Lymphocyte count,<br>× 10<sup>9</sup>/L* | 0.53(0.09-3.03) | 0.474 | .. | .. |
| Lactate dehydrogenase, U/L | 1.01(1.00-1.02) | 0.003 | .. | .. |
| ≤225 | 1 (ref) | .. | .. | .. |
| >225 | 6.15(0.66-57.60) | 0.111 | .. | .. |
| D-dimer, µg/mL | 9.30(1.85-46.74) | 0.007 | .. | .. |
| ≤0.5 | 1 (ref) | .. | .. | .. |
| >0.5 to ≤1 | .. | .. | .. | .. |
| >1 | 23.33(1.99-273.29) | 0.012 | .. | .. |
| Hypersensitive troponin I,<br>pg/mL | 1.00(0.99-1.01) | 0.411 | .. | .. |
| ≤34.2 | 1 (ref) | .. | .. | .. |
| >34.2 | 7.50(1.14-49.26) | 0.036 | .. | .. |
| **CT findings within the first week after symptom onset** |  |  |  |  |
| Total severity score* | 1.39(1.12-1.72) | 0.003 | 1.544(1.004-2.374) | 0.048 |
| <15 | 1 (ref) | .. | .. | .. |
| ≥15 | 35.00(3.32-368.57) | 0.003 | .. | .. |
| Number of involved lung<br>lobes | 1.366(1.003-1.860) | 0.048 | .. | .. |
| <5 | 1 (ref) | .. | .. | .. |
| =5 | 9.00(1.52-53.40) | 0.016 | .. | .. |

OR=odds ratio. \*Per 1 unit increase.

**Table S6.** Demographics and baseline characteristics of the subset of patients included in the analysis of CT within the first week after symptom onset.

|  | Total<br>(n=32) | Non-survivor<br>(n=11) | Survivor<br>(n=21) | p<br>value |
| --- | --- | --- | --- | --- |
| **Characteristics** |  |  |  |  |
| Age, years | 58(40-70) | 69(57-78) | 51(33-70) | 0.020 |
| <65 | 20 (62%) | 5 (45%) | 15 (71%) | 0.149 |
| ≥65 | 12 (38%) | 6 (55%) | 6 (29%) |  |
| Sex |  |  |  | 0.266 |
| Female | 13 (41%) | 3 (27%) | 10 (48%) |  |
| Male | 19 (59%) | 8 (73%) | 11 (52%) |  |
| Any Comorbidity | 13 (41%) | 6 (55%) | 7 (33%) | 0.246 |
| Diabetes | 5 (16%) | 2 (18%) | 3 (14%) | 0.773 |
| Hypertension | 9 (28%) | 5 (45%) | 4 (19%) | 0.115 |
| Coronary heart disease | 1 (3%) | 1 (9%) | 0 (0%) | 0.344 |
| Chronic obstructive<br>pulmonary disease | 1 (3%) | 1 (9%) | 0 (0%) | 0.344 |
| Malignancy | 0 (0%) | 0 (0%) | 0 (0%) | .. |
| Chronic liver disease | 1 (3%) | 0 (0%) | 1 (5%) | 1.000 |
| Other | 8 (25%) | 4 (36%) | 4 (19%) | 0.283 |
| Current smoker | 3 (9%) | 1 (9%) | 2 (10%) | 0.968 |
| **Symptoms and signs** |  |  |  |  |
| Fever | 30 (94%) | 10 (91%) | 20 (95%) | 0.631 |
| Highest temperature, °C | 38.6(38.0-39.0) | 38.6(37.6-39.0) | 38.5(38.0-39.0) | 0.611 |
| Chills | 7 (22%) | 3 (27%) | 4 (19%) | 0.593 |
| Cough | 23 (72%) | 9 (82%) | 14 (67%) | 0.365 |
| Sputum | 8 (25%) | 5 (45%) | 3 (14%) | 0.053 |
| Dyspnea | 16 (50%) | 5 (45%) | 11 (52%) | 0.710 |
| Hemoptysis | 2 (6%) | 1 (9%) | 1 (5%) | 0.631 |
| Chest pain | 2 (6%) | 1 (9%) | 1 (5%) | 0.631 |
| Headache | 3 (9%) | 1 (9%) | 2 (10%) | 0.968 |
| Fatigue | 8 (25%) | 2 (18%) | 6 (29%) | 0.519 |
| Nausea | 1 (3%) | 0 (0%) | 1 (5%) | 1.000 |
| Diarrhea | 5 (16%) | 3 (27%) | 2 (10%) | 0.189 |
| Myalgia | 7 (22%) | 3 (27%) | 4 (19%) | 0.593 |
| Systolic pressure, mm Hg | 129.0(114.0-144.0) | 144.0(125.0-171.0) | 126.0(112.0-139.0) | 0.074 |
| Heart rate, beats per minute | 95.0(81.0-110.0) | 103.0(86.0-111.0) | 86.0(80.0-109.0) | 0.367 |
| Respiratory rate | 20.0(20.0-24.0) | 24.0(20.0-25.0) | 20.0(20.0-22.0) | 0.022 |
| >20 breaths per min | 14 (44%) | 8 (73%) | 6 (29%) | 0.017 |
| Time from symptom onset<br>to hospital admission, days | 10.0(7.0-13.0) | 8.0(6.0-14.0) | 10.0(7.0-13.0) | 0.639 |

Data are median (IQR), n (%), or n/N (%), where N is the total number of patients with available data. $\chi^2$ test, Fisher's exact test, or Mann-Whitney U test were used to compare the values between survivors and non-survivors as appropriate.

**Table S7.** Laboratory findings of the subset of patients included in the analysis of CT within the first week after symptom onset.

|  | Normal range | Total<br>(n=32) | Non-survivor<br>(n=11) | Survivor<br>(n=21) | p value |
| --- | --- | --- | --- | --- | --- |
| White blood cell count, $\times 10^9/L$ | 4.00-10.00 | 5.2(4.2-8.3) | 9.1(6.2-11.8) | 4.7(4.1-5.4) | 0.000 |
| <4 |  | 4 (13%) | 0 (0%) | 4 (19%) | 0.002 |
| 4-10 |  | 23 (72%) | 6 (55%) | 17 (91%) |  |
| >10 |  | 5 (15%) | 5 (45%) | 0 (0%) |  |
| Neutrophil count, $\times 10^9/L$ | 1.80-6.30 | 3.5(2.8-6.9) | 8.0(3.5-11.3) | 2.9(2.7-4.3) | 0.000 |
| Lymphocyte count, $\times 10^9/L$ | 1.10-3.20 | 0.8(0.5-1.2) | 0.7(0.4-1.5) | 0.9(0.6-1.2) | 0.307 |
| <1.1 |  | 22 (69%) | 8 (73%) | 14 (67%) | 0.725 |
| $\geq 1.1$ | | 10 (31%) | 3 (27%) | 7 (33%) | |
| Hemoglobin, g/L | 130.0-175.0 | 126.5(120.0-140.0) | 120.0(110.0-135.0) | 129.0(122.5-141.5) | 0.113 |
| Platelet count, $\times 10^9/L$ | 125.0-350.0 | 162.0(124.0-242.0) | 124.0(87.0-242.0) | 175.0(133.8-287.0) | 0.066 |
| <125 |  | 8/31 (26%) | 6 (55%) | 2/20 (10%) | 0.007 |
| $\geq 125$ | | 23/31 (74%) | 5 (45%) | 18/20 (90%) | |
| Lactate dehydrogenase, U/L | 135-225 | 328.0(197.5-541.8) | 570.0(440.0-671.0) | 261.0(194.5-379.5) | 0.001 |
| $\leq 225$ | | 9 (28%) | 1 (9%) | 8 (38%) | 0.083 |
| >225 |  | 23 (72%) | 10 (91%) | 13 (62%) |  |
| D-dimer, $\mu g/mL$ | $\leq 0.5$ | 0.9(0.5-2.0) | 2.3(1.6-22.0) | 0.6(0.4-0.9) | 0.000 |
| $\leq 0.5$ | | 8 (25%) | 1 (9%) | 7 (33%) | 0.000 |
| >0.5 to $\leq 1$ | | 11 (34%) | 0 (0%) | 11 (52%) | |
| >1 |  | 13 (41%) | 10 (91%) | 3 (15%) |  |
| Prothrombin time, s | 11.5-14.5 | 14.8(13.8-15.4) | 15.5(14.8-17.7) | 14.3(13.4-15.0) | 0.005 |
| <14.5 |  | 14 (44%) | 2 (18%) | 12 (57%) | 0.035 |
| ≥14.5 |  | 18 (56%) | 9 (82%) | 9 (43%) |  |
| International Normalized Ratio, INR | 0.80-1.20 | 1.15(1.05-1.20) | 1.21(1.14-1.45) | 1.09(1.01-1.17) | 0.005 |
| Hypersensitive troponin I, pg/mL | ≤34.2 | 11.0(4.2-27.0) | 23.1(13.0-204.7) | 6.0(2.3-13.2) | 0.005 |
| ≤34.2 |  | 24/31 (77%) | 6 (55%) | 18/20 (90%) | 0.024 |
| >34.2 |  | 7/31 (23%) | 5 (45%) | 2/20 (10%) |  |
| NT-proB-type Natriuretic Peptide (BNP), pg/mL | <241 | 192.0(71.0-702.5) | 777.0(348.5-2847.0) | 90.5(58.8-194.5) | 0.000 |
| <241 |  | 17/30 (57%) | 0 (0%) | 17/20 (85%) | 0.000 |
| ≥241 |  | 13/30 (43%) | 10 (100%) | 3/20 (15%) |  |
| Albumin, g/L | 35-52 | 33.4(30.1-39.5) | 31.5(28.2-34.2) | 36.0(32.2-39.9) | 0.034 |
| <35 |  | 18 (56%) | 9 (82%) | 9 (43%) | 0.035 |
| ≥35 |  | 14 (44%) | 2 (18%) | 12 (57%) |  |
| Alanine aminotransferase, U/L | ≤41 | 19.5(13.0-28.5) | 17.0(14.0-29.0) | 20.0(12.0-27.5) | 0.725 |
| ≤41 |  | 28 (87%) | 10 (91%) | 18 (86%) | 0.673 |
| >41 |  | 4 (13%) | 1 (9%) | 3 (14%) |  |
| Aspartate aminotransferase, U/L | ≤40 | 30.0(19.3-43.0) | 34.0(24.0-54.0) | 24.0(19.0-36.0) | 0.271 |
| ≤40 |  | 24 (75%) | 7 (64%) | 17 (81%) | 0.283 |
| >40 |  | 8 (25%) | 4 (36%) | 4 (19%) |  |
| Total bilirubin, μmol/L | ≤26 | 7.9(6.2-10.3) | 7.5(6.6-9.8) | 8.1(6.0-10.9) | 1.000 |
| Creatinine, μmol/L | 59-104 | 68.0(61.3-87.8) | 100.0(62.0-164.0) | 67.0(59.5-76.0) | 0.088 |
| ≤104 |  | 27 (84%) | 6 (55%) | 21 (100%) | 0.002 |
| >104 |  | 5 (16%) | 5 (45%) | 0 (0%) |  |
| Blood urea nitrogen, mmol/L | 3.6-9.5 | 4.7(3.3-8.4) | 9.2(5.4-13.7) | 4.0(3.3-5.2) | 0.008 |
| ≤9.5 |  | 26 (81%) | 6 (55%) | 20 (95%) | 0.005 |
| >9.5 |  | 6 (19%) | 5 (45%) | 1 (5%) |  |
| Potassium, mmol/L | 3.50-5.10 | 4.1(3.5-4.5) | 4.4(3.6-5.3) | 4.0(3.5-4.3) | 0.208 |
| <3.5 |  | 7 (22%) | 2 (18%) | 5 (24%) | 0.042 |
| 3.5-5.1 |  | 22 (69%) | 6 (55%) | 16 (76%) |  |
| >5.1 |  | 3 (9%) | 3 (27%) | 0 (0%) |  |
| Sodium, mmol/L | 136-145 | 139.0(136.4-141.9) | 138.7(137.2-142.6) | 139.1(135.6-141.7) | 0.938 |
| <136 |  | 7 (22%) | 2 (18%) | 5 (24%) | 0.363 |
| 136-145 |  | 24 (75%) | 8 (73%) | 16 (76%) |  |
| >145 |  | 1 (3%) | 1 (9%) | 0 (0%) |  |
| Calcium, mmol/L | 2.20-2.55 | 2.1(2.0-2.2) | 2.0(1.9-2.2) | 2.1(2.0-2.2) | 0.081 |
| <2.2 |  | 25 (78%) | 10 (91%) | 15 (71%) | 0.205 |
| ≥2.2 |  | 7 (22%) | 1 (9%) | 6 (29%) |  |
| Procalcitonin, ng/mL | <0.05 | 0.05(0.03-0.18) | 0.32(0.11-0.60) | 0.04(0.02-0.06) | 0.000 |
| <0.05 |  | 17 (53%) | 1 (9%) | 16 (76%) | 0.000 |
| ≥0.05 |  | 15 (47%) | 10 (91%) | 5 (24%) |  |
| High-sensitivity C-reactive Protein (hs-CRP), mg/L | <1 | 52.2(9.6-114.7) | 102.1(51.3-194.8) | 27.0(5.5-71.1) | 0.009 |
| <3 |  | 4 (13%) | 0 (0%) | 4 (19%) | 0.272 |
| ≥3 |  | 28 (87%) | 11 (100%) | 17 (81%) |  |
| IL-1β, pg/ml | <5 | 4.9(4.7-4.9) | 4.9(4.2-4.9) | 4.9(4.7-4.9) | 0.652 |
| IL-2R, U/ml | 223-710 | 514.0(292.5-744.5) | 1076.5(671.8-1699.5) | 454.5(270.3-563.0) | 0.005 |
| IL-6, pg/ml | <7 | 4.5(1.4-22.6) | 65.1(11.3-154.1) | 3.3(1.4-16.7) | 0.019 |
| IL-8, pg/ml | <62 | 10.2(6.1-18.9) | 27.6(14.1-64.9) | 9.4(5.7-15.9) | 0.010 |
| IL-10, pg/ml | <9.1 | 4.9(4.0-4.9) | 8.3(4.9-17.0) | 4.9(4.0-4.9) | 0.081 |
| TNF-α, pg/ml | <8.1 | 7.0(4.9-9.6) | 21.3(13.2-28.9) | 5.7(3.8-7.9) | 0.000 |

Data are median (IQR), n (%), or n/N (%), where N is the total number of patients with available data. $\chi^2$ test, Fisher's exact test, or Mann-Whitney U test were used to compare the values between survivors and non-survivors as appropriate.

**Figure S1.**
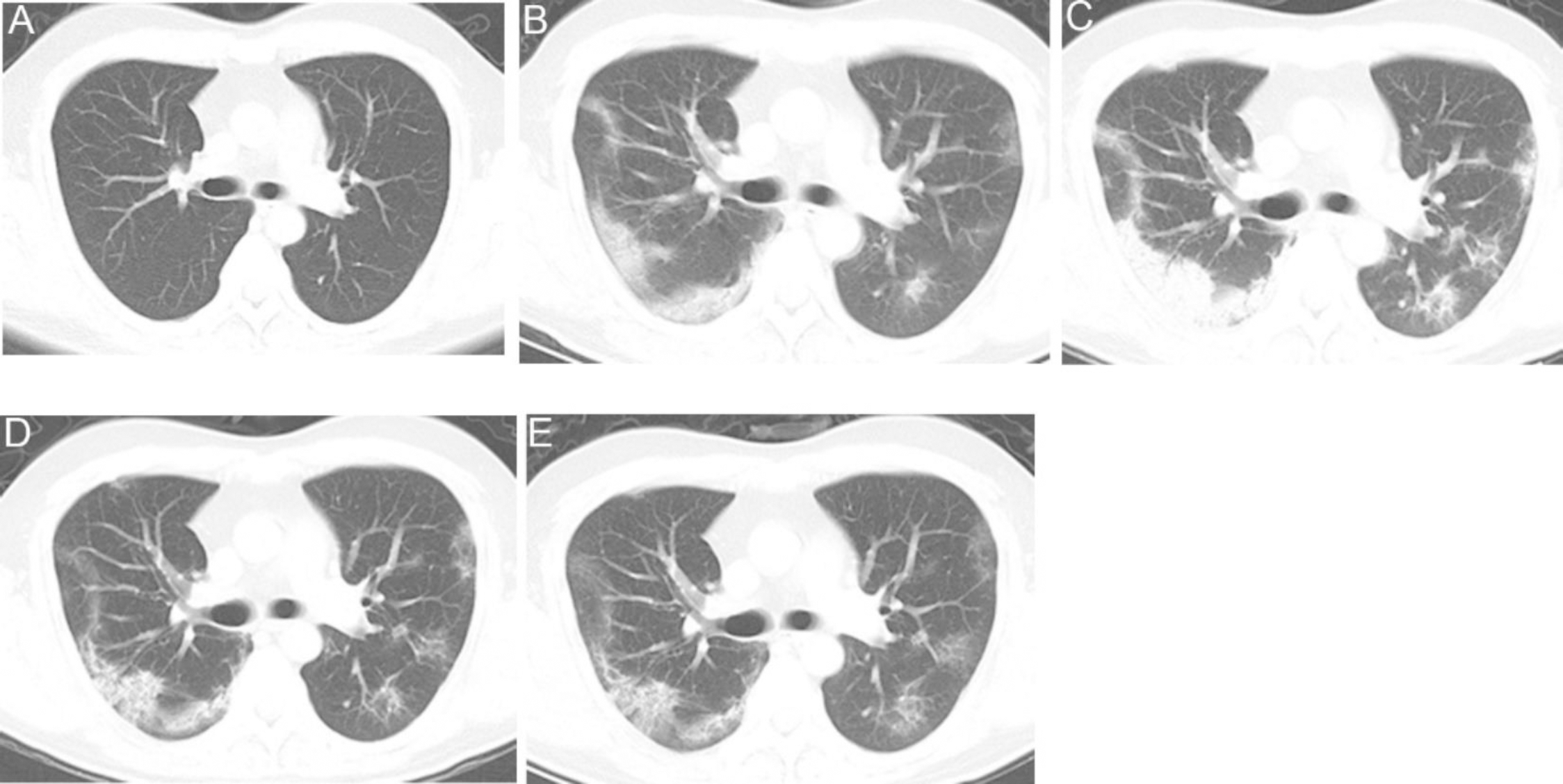
Transverse CT images of a 38-year-old man with Covid-19. A, Normal CT images on the day when the patient had initial symptoms (day 1). B, day 9 after symptom onset, bilateral and peripheral ground-grass opacity associated smooth interlobular and intralobular septal thickening (crazy-paving pattern). C, day 15 after symptom onset, peripheral predominant consolidation pattern with air bronchograms in right upper and lower lobes. D, day 23 after symptom onset, previous opacifications were dissipated into ground-grass opacities and irregular interlobular and intralobular septal thickening (reticulation pattern). E, day 30 after symptom onset, further resolution of the lesions, ground-glass opacities and reticulation patterns remained.

**Figure S2.**
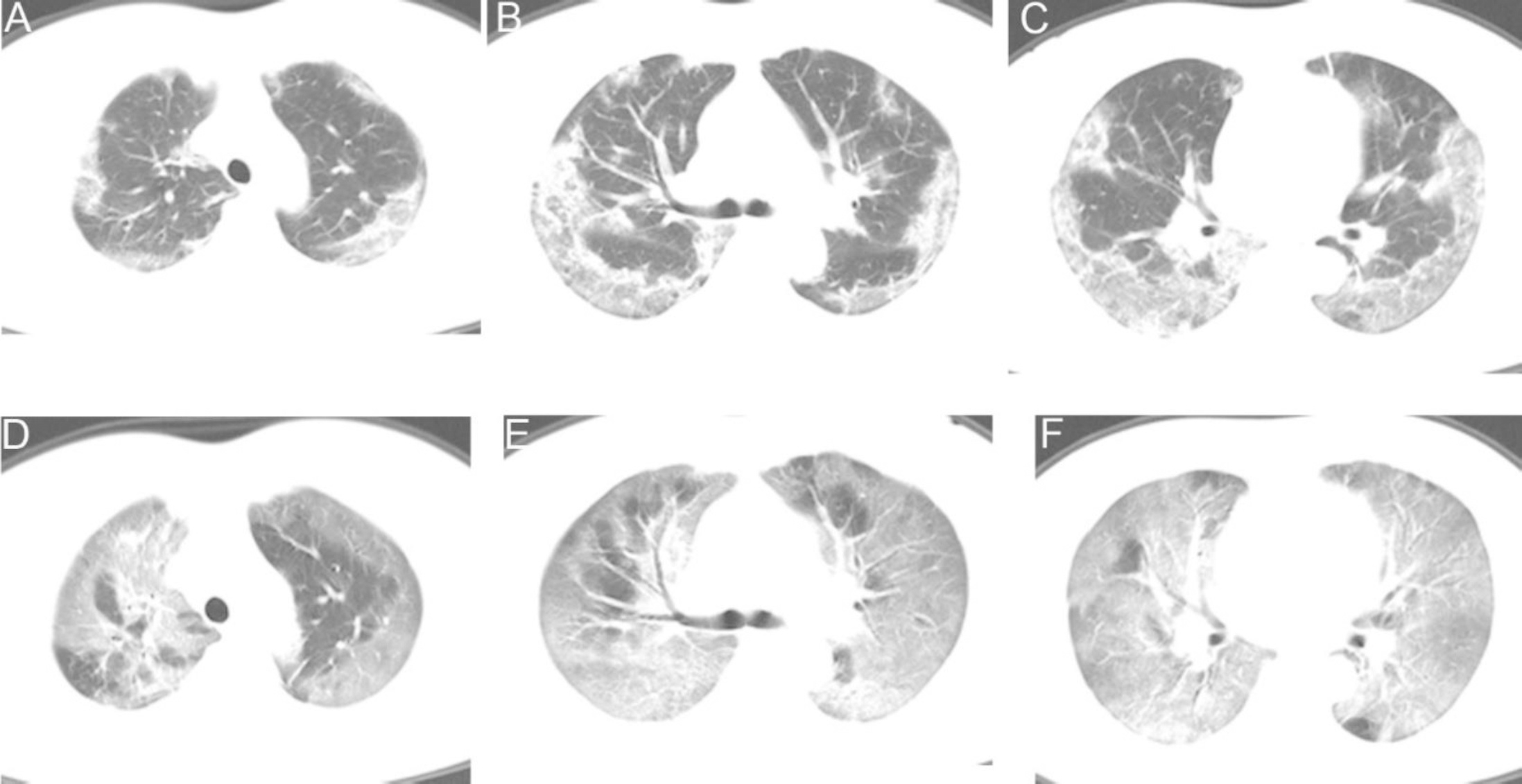
Transverse CT images of a 41-year-old woman with Covid-19. A-C, day 6 after symptom onset, multifocal consolidations and ground-glass opacities affecting the bilateral, subpleural lung parenchyma associated with crazy-paving pattern. D-F, day 10 after symptom onset, bilateral extensive ground-glass opacities, involving nearly the entire lower lobes and right middle lobe, and most of the upper lobes, giving a white lung appearance, with air bronchograms and crazy-paving pattern. The patient died 8 days after this scan.

## Author Contributions

KL and GZ conceptualized the study design. KL, SC, DC, YF, CC collected demographic, clinical, and laboratory data. ZW and NW interpreted the images of CT scans. KL, DC, YF and GZ analysed the data. KL and GZ interpreted the results. GZ wrote the manuscript with all authors providing feedback for revision. All authors read and approved the final report.

## Acknowledgement

We are very grateful to all members of the medical, nursing, and support staffs of the quarantine unit at the Sino-French branch of Tongji Hospital for their support. We are very grateful to David J. Erle (University of California San Francisco) for the critical review of the manuscript.

